# Pregnancy-related risk and birth setting trends: Insights from Indonesian Demographic Health Survey data

**DOI:** 10.1101/2025.01.19.25320813

**Authors:** Kai Hodgkin, Grace Joshy, Kamalini Lokuge

**Affiliations:** National Centre for Epidemiology and Population Health, Australian National University; Discipline of Midwifery, University of Canberra Corresponding Author

**Author notes:** Kai Hodgkin National Centre for Epidemiology and Population Health College of Health and Medicine Australian National University 62 Mills Road Acton ACT 2600 Australia.

**Keywords:** Indonesia, Pregnancy, Birth Setting, High Risk Pregnancy, Referral and Consultation

## Abstract

**Background:** Most maternal and neonatal deaths occur in low- and middle-income countries and are largely preventable with quality care. In Indonesia, 98% of pregnant people receive antenatal care and birth commonly occurs in community settings or hospitals. Outside of high-income countries, few studies identify where women with pregnancy-related risk factors give birth. Accurate identification of pregnancy risk factors and referral to an appropriate birth setting is considered an essential element of quality antenatal care, though its efficacy in Indonesia is unknown.

**Methods:** This study aimed to identify suitable indicators of pregnancy risk factors for Indonesia and examine population-level patterns in pregnancy-related risk, care and appropriateness of birth setting. Risk factors in pregnancy based on internationally relevant referral guidelines were identified from literature search and mapped to available indicators. Using self-reported data from three waves of the Indonesian Demographic Health Survey (2007, 2012, 2017), a representative survey of women aged 15-49 years, we examined receipt of maternity care, prevalence of pregnancy risk factors and time trends in birth setting, overall and by presence of risk factors.

**Results:** In the weighted sample (n=43,846), one quarter of women reported pregnancy risk factors. From 2002-2017, numbers of births in hospitals have doubled and births at home have halved. However, the proportions of women with pregnancy risk in each setting remains largely unchanged.

**Discussion:** These findings identify opportunities for shifting care in Indonesia to ensure women are receiving the appropriate level of care at birth.

## Introduction

Ending neonatal and maternal mortality remains a global challenge still to be met. In 2019 the world recorded 2 440 464 neonatal deaths and in 2017 there were 295 000 maternal deaths ^1^, the vast majority in low and middle income countries. Most of these deaths, as well as the additional burden of morbidity issues are preventable with appropriate and timely birth care ^2^. While improving, the world still has a long way to go in strengthening health care as an important aspect to ending these deaths.

In 2014 an influential Lancet series published a framework for quality maternal and newborn care ^3^. The framework is based on 461 systematic reviews and identifies that the best care provision is based on a tiered system where all women and babies have access to skilled, preventative care and health promotion alongside first line management of complications. The second tier is medical obstetric and neonatal services for the management of serious complications for those who need it. They argue that midwifery is best placed to provide this first tier of care for all and that the promotion of normal reproductive processes has an important role to play in reducing the iatrogenic effects of growing over-intervention in maternity care.

This framework, and indeed most functioning maternity care systems depend on the accurate and timely identification and referral of those who require management of complex interventions ^4^.

However, many barriers to accessing the ideal level of care complicate women’s use of health facilities, and have been well described by the “delays model” ^5,6^. These delays include a delay in decision to seek care, delays in reaching a health facility and delays in provision of appropriate, quality care. Therefore, there is a clear need to identify where women in need of different levels of care are giving birth in order to then assess if there are delays in access which need to be overcome.

There is no consensus on how to define risk during pregnancy which deems a woman at increased risk of obstetric complications, though such screening and referral is seen as essential and occurs continuously in antenatal care the world over ^7^. There are some complications which clearly indicate need for hospital care such as placenta previa which will result in serious harm and likely death to a woman and her baby if a caesarean birth is not performed ^8^. Other factors are more grey or controversial, however. For example, previous caesarean section is considered a risk requiring increased monitoring provided in a hospital in the UK ^9^, yet in Canada is considered a low risk and therefore acceptable for home birth ^10^. Additionally, some complications, such as malaria are extremely rare and therefore not screened for in settings such as Australia ^11^, yet are far more common and should be screened for in other settings like Indonesia ^12^.

Despite the clear need to identify and triage care appropriately for women with complications in pregnancy, studies in low and middle income countries rarely, if at all, consider pregnancy or birth complications when assessing outcomes by birth settings ^13^. Therefore, there is no clear understanding of how well triaging to emergency obstetric health services on the basis of pregnancy and birth complications is occurring in low and middle income countries. While this type of data is routinely collected in high income countries, many low and middle income countries do not have national pregnancy and birth datasets and therefore detail of pregnancy and birth care associated with complications is lacking ^14^. This kind of data is commonly only available at a hospital or health district level.

This article fills this gap by identifying in a population level dataset (Demographic Health Survey) pregnancy complications occurring prior to onset of labour that meet guidelines for referral to higher level services due to increased risk of adverse outcomes. We then describe where these women with and without pregnancy risks gave birth which indicates whether appropriate triaging has occurred.

Indonesia is the focus of this study because of both the availability of pregnancy risk data and a health system which largely fits with Renfrew’s Framework of skilled pregnancy care for all women with access to obstetric and neonatal care for those that need it. Indonesia’s maternity care system is based in community midwifery care for all via the *bidan di desa* (village midwife) program and 98% of women receive antenatal care ^15^. The health insurance program reduces the barrier of cost to accessing medical care for those that need it and the numbers of women who give birth in a hospital setting is steadily rising. Finally, the Indonesian Demographic Health Survey asks women whether they experienced complications in pregnancy.

Despite the high levels of perinatal care, mortality remains high in Indonesia. The neonatal mortality rate is 11 per 1000 births which is higher than the average of the region^16^. Indonesia is the fourth most populous country in the world, meaning this represents a high burden of the world’s neonatal deaths. The leading causes of neonatal mortality are premature birth and intrapartum events^17^ and preventing these deaths is dependent on appropriate care at birth. Therefore it is important to evaluate whether women are receiving the appropriate level of care at birth for their pregnancy-related risk, or if more could be done to ensure quality care.

### Aim

We developed two specific aims for this project:

1. Outline a set of indicators for use in Indonesia which define women with pregnancies at risk of obstetric complications during birth, and well women with lower risk of obstetric complications during birth utilising Indonesian Demographic Health survey data variables.
2. Determine trends in birth settings overall, and for women with and without pregnancy risk in Indonesia.

## Methods

### Reporting

The Strengthening the Reporting of Observational Studies in Epidemiology (STROBE) Checklist for cross-sectional studies was used ^18^. The checklist is available in Appendix 1.

### Data

Cross sectional Demographic Health Survey (DHS) data from Indonesia was utilised for this investigation including the most recent three waves of the survey: 2007, 2012, 2017. DHS is a USAid funded international initiative which focuses on collecting high quality, representative survey data in conjunction with lower and middle income countries. Each wave of the survey involves a centrally developed questionnaire which is then modified, translated and administered by each local country involved. The dataset is de-identified, coded, cleaned and released publicly on the DHS website for those who register. The process of participant selection, data collection and documentation is well documented elsewhere ^15^.

Each of the three waves of the DHS survey were first cleaned to address differences in coding conventions for the same question between the waves, and then merged to form a single dataset.

### Population

The population was defined as women aged 15-49, interviewed in the 2007, 2012 or 2017 Indonesian DHS waves, who had given birth to their most recent baby (or babies in the case of multiples) within five years of the survey. In 2007, the survey was only conducted among married women, while in 2012 and 2017, unmarried women were also included.

### Ethics

[removed for blind review]

### Defining increased risk of requiring obstetric intervention

We aimed to identify a definition of increased risk of obstetric complications in pregnancy which is applicable in Indonesia and from an international organisation such as the World Health Organisation. To do this, we searched the internet for relevant care guidelines which included a list of risk factors which would result in the recommendation of referral to a setting with emergency obstetric (and/or neonatal) care and facilities. These risk factors were then compiled and organised by domain (maternal, neonatal and obstetric history). The Indonesian DHS dataset was then searched for an equivalent or proxy variable for each of these factors which could be utilised in this analysis.

We then undertook an analysis of place of birth by the number of risk factors a woman self-reported. This was based on an assumption that the greater the number of risk factors a woman experiences in her pregnancy the greater the risk of complications at birth, and therefore the more important it would be that the woman gives birth in a setting with complex care available (tier two from the Renfrew Framework ^3^).

### Categorising birth settings

Indonesia has a range of types of health facilities where women birth ^19^. Across each of the three waves of the Indonesian DHS these were defined differently so existing categories could not be used. Additionally, some categories contained too few births to result in useful findings. For the purposes of this study, birth settings were grouped into categories aiming to approximate the level of tier two care ^20^ available: Hospitals (including public, private and maternity hospitals); other clinics (these include Puskesmas, Posyandu, PolIndes, private nurse, GP and obstetric practices); midwifery clinics (including government funded and private practice clinics); and homebirth (including another person’s home) where a health professional (mostly a midwife) is present. The final category is all births not attended by a health professional (many were attended by a dukun bayi or traditional birth attendant), 98 per cent of which occur at home and also includes most births in the ‘other’ category which (while unclear) likely involves babies born en route to a health facility or at un- or under-staffed clinics.

While the categories do not explicitly equate to the types of services available in each setting, we assume that at the majority of both public and private hospitals, Comprehensive Emergency Obstetric and Neonatal Care (CEmONC) facilities will be available while clinics (including midwifery clinics) might have access to some, if not all Basic Emergency Obstetric and Neonatal Care requirements (BEmONC). BEmONC facilities are usually able to provide life-saving medications, intravenous therapy, basic neonatal resuscitation and assisted birth ^20^. CEmONC facilities can provide the same as BEmONC in addition to emergency surgery (caesarean section) and specialist obstetric and neonatal care. Homes are unlikely to have any of these, though if health professionals are in attendance, they likely bring assessment tools, sterile birth kits, some medications, intravenous therapy and basic resuscitation equipment.

### Analyses

Weighted proportions were estimated, applying DHS-provided weights for each wave. In this population the weighting includes correcting for non-response and oversampling of women in rural households. Initial cross-tabulations were undertaken ensuring data accuracy. Analyses were conducted using Stata 17^21^. Chi-square test was used to determine significance of proportions at the 0.001 significance level.

## Results

### Definition of pregnancy risk

Searches for care guidelines containing lists of risk factors which would define a pregnant person at greater risk of obstetric complication resulted in two potential sources: the Indonesian *Buku Saku: Pelayanan Kesehatan Ibu di Fasilitas Kesehatan Dasar dan Rujuka*(*nHandbook: Healthcare of mothers in basic health facilities and referral*) ^12^ and the WHO and UNICEF’s *Pregnancy, childbirth, postpartum and newborn care: a guide for essential practice* ^22^.

The Indonesian *Buku Saku: Pelayanan Kesehatan Ibu di Fasilitas Kesehatan Dasar dan Rujukan* is aimed at providing guidance for health professionals in primary health settings in Indonesia and was published in partnership with the World Health Organisation ^12^. It offers a list of pregnancy factors which are used to recommend a woman be transferred to a tertiary institution before birth. The second source used is the WHO and UNICEF’s *Pregnancy, childbirth, postpartum and newborn care:*

*a guide for essential practice* ^22^. This document advises health professionals to recommend that all women birth in a health facility, and provides a list of reasons that a woman should birth in a better equipped (referral level) setting. Neither provides supporting evidence for their recommendations as is expected for WHO guidelines ^23^, meaning that the quality of the recommendations cannot be determined. Table 1 lists the risk factors defined in both sources which have reasonable proxies in the Indonesian DHS dataset. A binary variable was created which categorised women as either having or not having any of the pregnancy complications listed in Table 1.

**Table 1.**
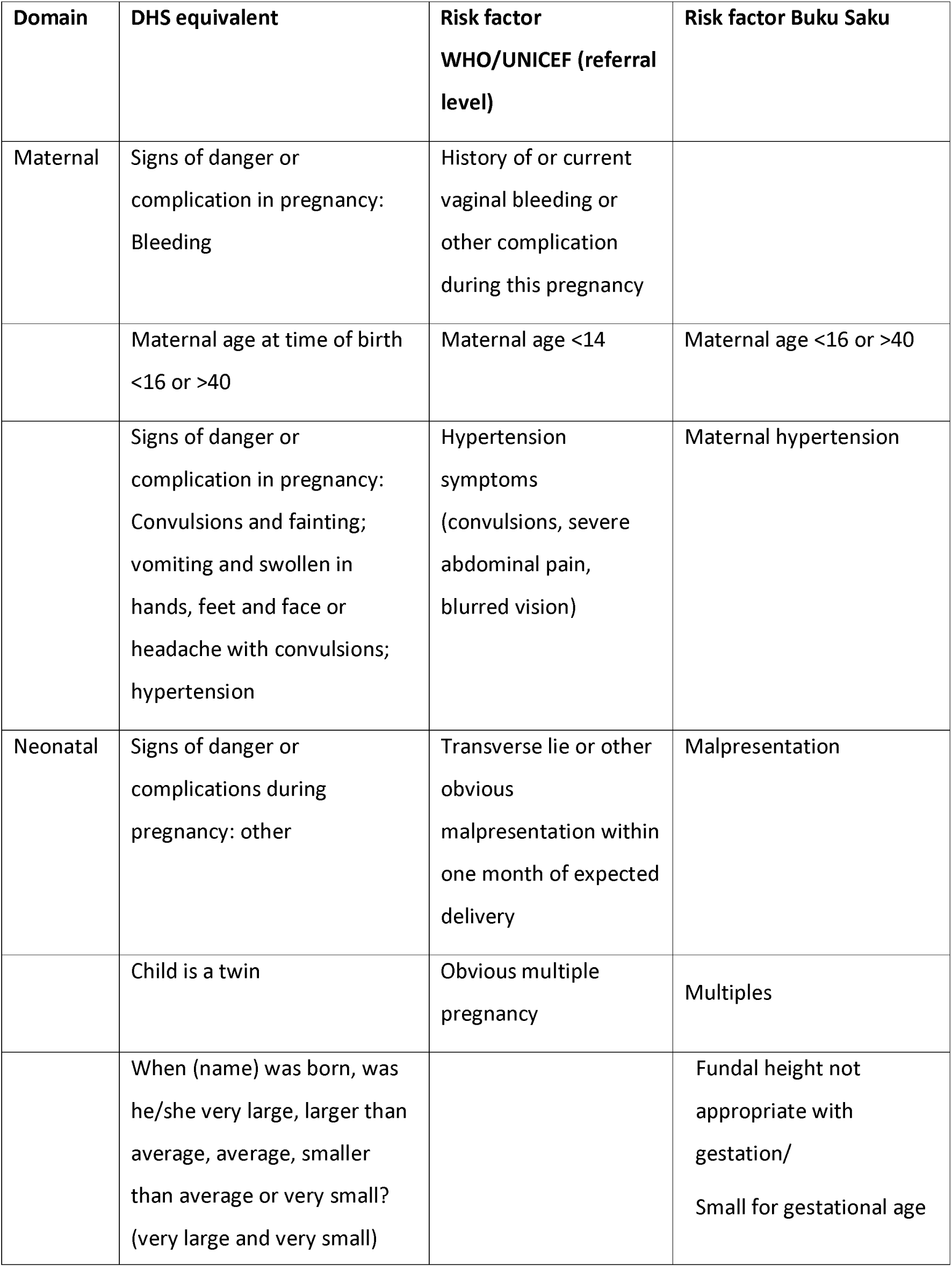
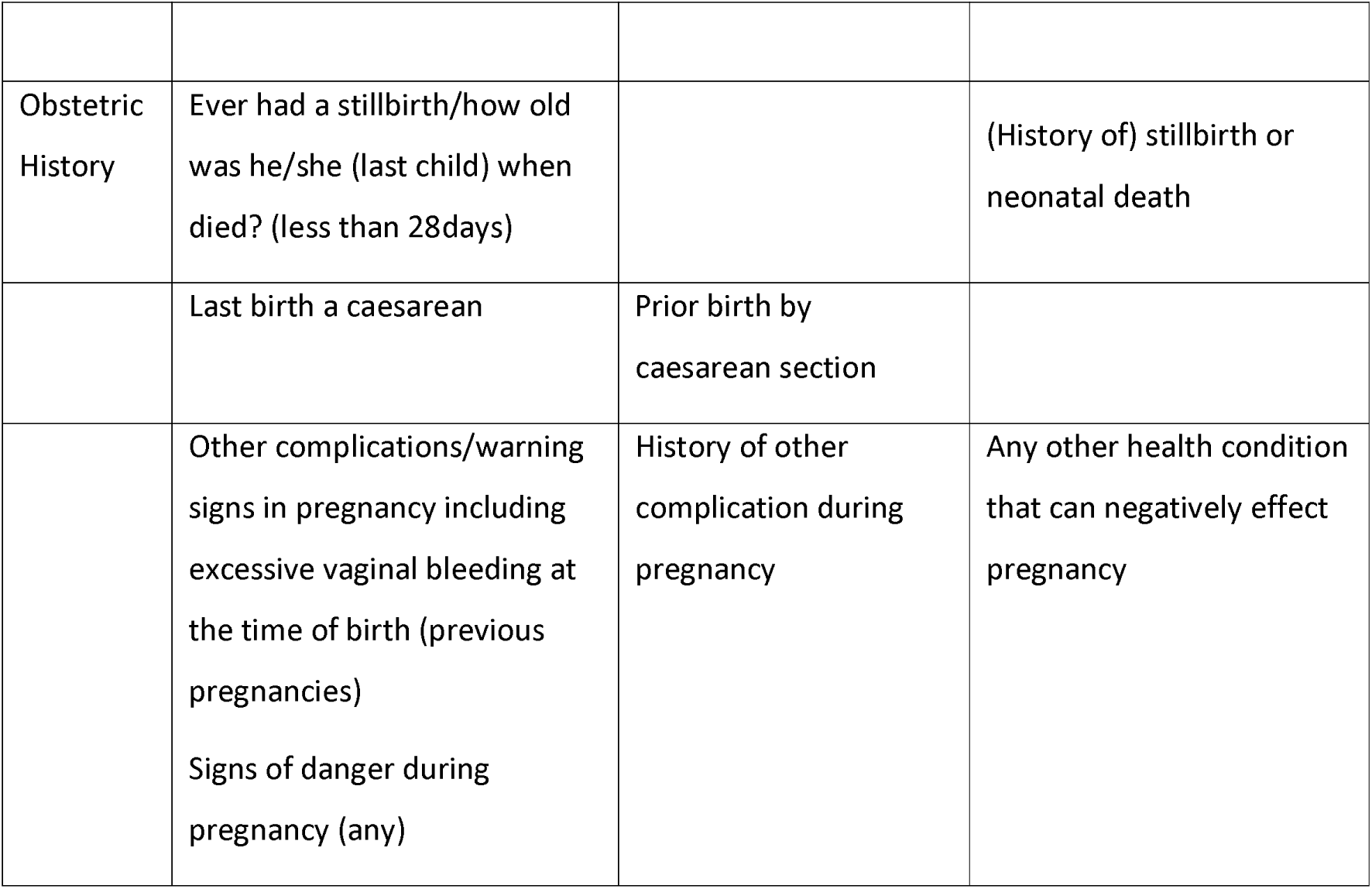
Definition of pregnancy risk factors which place a woman at higher risk of obstetric complications and recommend referral to tier two care (specialist obstetric/neonatal care) here

Additional risk factors were listed by the WHO/UNICEF or the Buku Saku but do not have DHS equivalents and therefore were not included in the definition of risk. The DHS category of ‘other complications/warning signs in pregnancy’ may include some of these but they were not specifically asked about in the survey. These include:

- Severe febrile disease or upper urinary tract infection
- Prior delivery by forceps or vacuum
- Documented history of third degree perineal tear
- Tubal ligation or IUD desired immediately after delivery
- Possible pneumonia/Lung disease
- HIV-infected woman
- Severe anaemia
- History of convulsions in previous pregnancy.

### Population Demographic Factors

Data were extracted from the Indonesian DHS dataset to define the demographic characteristics and are listed in Table 2. In total, 45,953 women from the total of 128,129 met the criteria of having given birth to their most recent baby within five years prior to the survey. Once weighting was applied, the total was 43,846 women. Table 2 describes the demographics of women included.

**Table 2:**
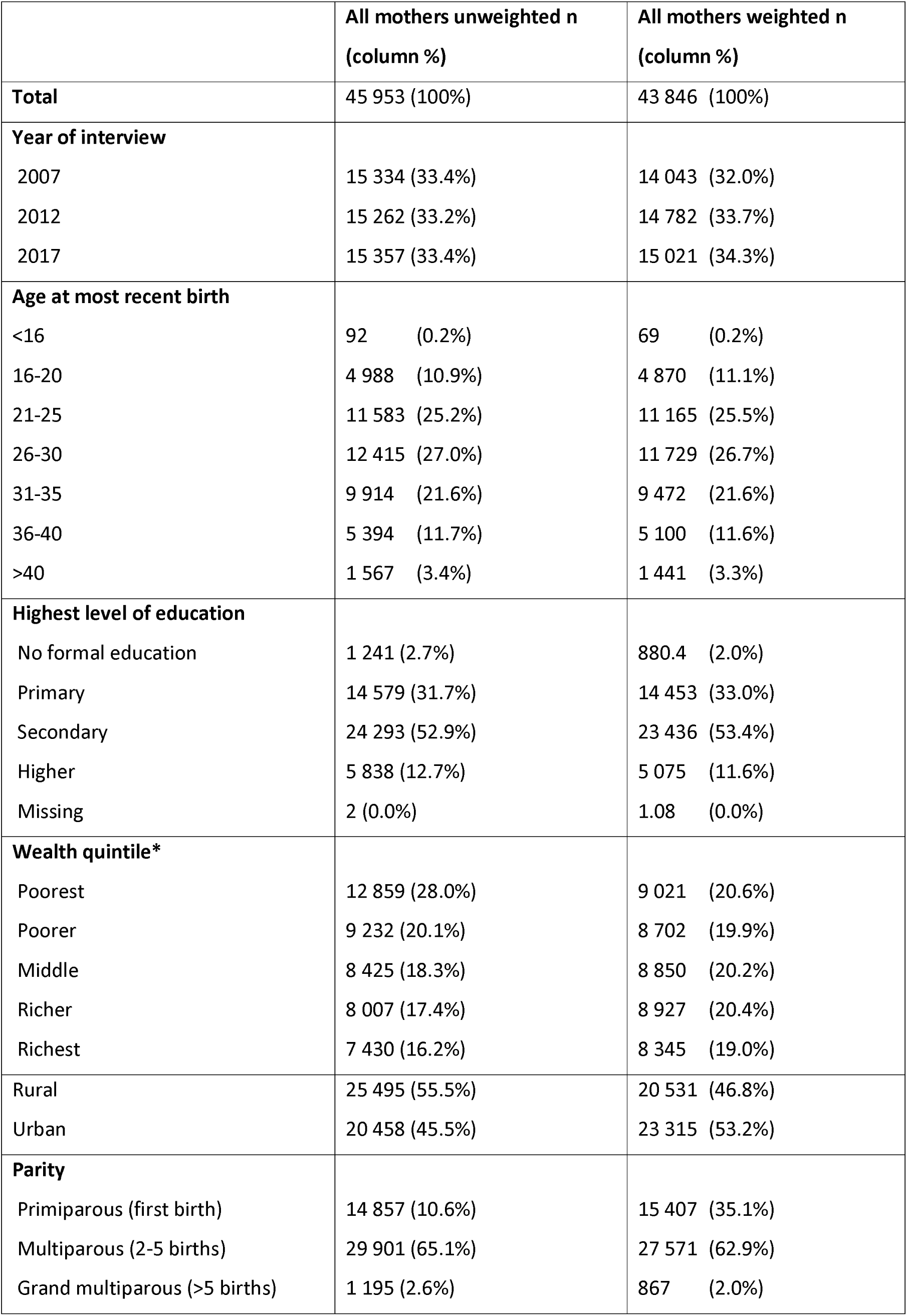
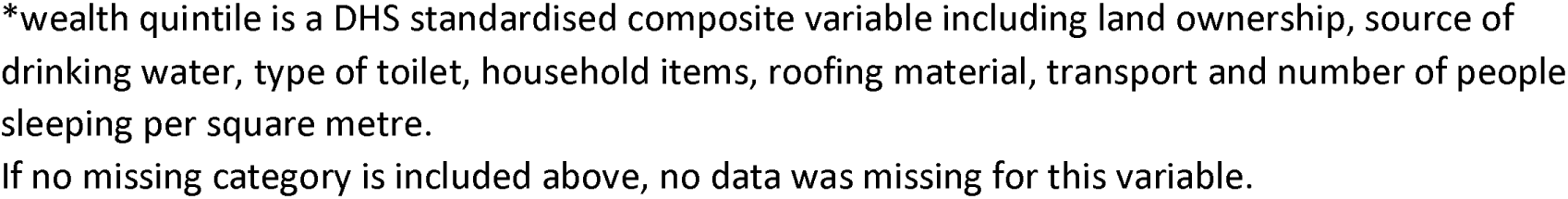
Population demographic factors here

|  | All mothers unweighted n<br>(column %) | All mothers weighted n<br>(column %) |
| --- | --- | --- |
| **Total** | 45 953 (100%) | 43 846 (100%) |
| **Year of interview** |  |  |
| 2007 | 15 334 (33.4%) | 14 043 (32.0%) |
| 2012 | 15 262 (33.2%) | 14 782 (33.7%) |
| 2017 | 15 357 (33.4%) | 15 021 (34.3%) |
| **Age at most recent birth** |  |  |
| <16 | 92 (0.2%) | 69 (0.2%) |
| 16-20 | 4 988 (10.9%) | 4 870 (11.1%) |
| 21-25 | 11 583 (25.2%) | 11 165 (25.5%) |
| 26-30 | 12 415 (27.0%) | 11 729 (26.7%) |
| 31-35 | 9 914 (21.6%) | 9 472 (21.6%) |
| 36-40 | 5 394 (11.7%) | 5 100 (11.6%) |
| >40 | 1 567 (3.4%) | 1 441 (3.3%) |
| **Highest level of education** |  |  |
| No formal education | 1 241 (2.7%) | 880.4 (2.0%) |
| Primary | 14 579 (31.7%) | 14 453 (33.0%) |
| Secondary | 24 293 (52.9%) | 23 436 (53.4%) |
| Higher | 5 838 (12.7%) | 5 075 (11.6%) |
| Missing | 2 (0.0%) | 1.08 (0.0%) |
| **Wealth quintile\*** |  |  |
| Poorest | 12 859 (28.0%) | 9 021 (20.6%) |
| Poorer | 9 232 (20.1%) | 8 702 (19.9%) |
| Middle | 8 425 (18.3%) | 8 850 (20.2%) |
| Richer | 8 007 (17.4%) | 8 927 (20.4%) |
| Richest | 7 430 (16.2%) | 8 345 (19.0%) |
| Rural | 25 495 (55.5%) | 20 531 (46.8%) |
| Urban | 20 458 (45.5%) | 23 315 (53.2%) |
| **Parity** |  |  |
| Primiparous (first birth) | 14 857 (10.6%) | 15 407 (35.1%) |
| Multiparous (2-5 births) | 29 901 (65.1%) | 27 571 (62.9%) |
| Grand multiparous (>5 births) | 1 195 (2.6%) | 867 (2.0%) |

\*wealth quintile is a DHS standardised composite variable including land ownership, source of drinking water, type of toilet, household items, roofing material, transport and number of people sleeping per square metre.
If no missing category is included above, no data was missing for this variable.

Table 3 shows that while around 95% of women in the survey reported receiving some antenatal care, only 58% received the currently recommend adequate care (8 visits or more with a health professional during their most recent pregnancy). Nearly three quarters of all women reported no pregnancy risk factors. Of the 10693 women who did report one or more risk factors, 3689 gave birth in a hospital (35%). The most common birth setting overall was a midwifery clinic with around 29% of women, followed by a hospital where 24% of women reported giving birth. 19% gave birth at home with a health professional and 16% gave birth without a health professional present, totalling 35% who gave birth outside of any health facility.

**Table 3:** Maternity Care and Reported Pregnancy Risk Factors – All mothers weighted (column %) here

|  | Midwifery Clinic | Other Clinic | Hospital | Homebirth | No health professional present | Missing | Total |
| --- | --- | --- | --- | --- | --- | --- | --- |
| **Total** | 12581<br>(100.0%) | 5229<br>(100.0%) | 10460<br>(100.0%) | 8391<br>(100.0%) | 7075<br>(100.0%) | 112<br>(100.0%) | 43846<br>(100.0%) |
| **Antenatal Care\*\*** |  |  |  |  |  |  |  |
| 0 visits with a health professional | 97 (0.8%) | 49 (0.9%) | 93 (0.9%) | 263 (3.1%) | 1377 (19.5%) | 89 (79.5%) | 1968 (4.5%) |
| <4 visits (inadequate care) | 633 (5.0%) | 279 (5.3%) | 434 (4.1%) | 1054 (12.6%) | 1513 (21.4%) | 3 (3.1%) | 3915 (8.9%) |
| 4-7 visits (adequate for pre-2016 WHO recommendation, inadequate for current WHO recommendation) | 3181 (25.3%) | 1476 (28.2%) | 2415 (23.1%) | 3220 (38.4%) | 2234 (31.6%) | 2 (1.9%) | 12528 (28.6%) |
| 8+ visits (adequate care currently recommended by WHO) | 8607 (68.4%) | 3402 (65.1%) | 7475 (71.5%) | 3787 (45.1%) | 1909 (27.0%) | 17 (15.3%) | 25198 (57.5%) |
| Missing | 63 (0.5%) | 24 (0.5%) | 42 (0.4%) | 67 (0.8%) | 41 (0.6%) | 0 (0.2%) | 237 (0.5%) |
| **Reported risk factors** |  |  |  |  |  |  |  |
| Any risk factors | 2587 (20.6%) | 1277 (24.4%) | 3689 (35.3%) | 1658 (19.8%) | 1457 (20.6%) | 25 (22.3%) | 10693 (24.4%) |
| No risk factors | 9993 (79.4%) | 3952 (75.6%) | 6770 (64.7%) | 6733 (80.2%) | 5618 (79.4%) | 87 (77.7%) | 33153 (75.6%) |
| **Number of risk factors** |  |  |  |  |  |  |  |
| 0 | 9993 (79.4%) | 3952 (75.6%) | 6770 (64.7%) | 6733 (80.2%) | 5618 (79.4%) | 87 (77.6%) | 33153 (75.6%) |
| 1 | 2363 (18.8%) | 1162 (22.2%) | 3094 (29.6%) | 1502 (17.9%) | 1328 (18.8%) | 24 (21.5%) | 9473 (21.6%) |
| 2 | 208 (1.7%) | 109 (2.1%) | 519 (5.0%) | 140 (1.7%) | 121 (1.7%) | 1 (0.8%) | 1097 (2.5%) |
| 3 | 15 (0.1%) | 3 (0.1%) | 61 (0.6%) | 12 (0.1%) | 6 (0.1%) | 0 (0.1%) | 97 (0.2%) |
| 4 | 1 (0.0%) | 3 (0.1%) | 16 (0.2%) | 4 (0.1%) | 2 (0.0%) | 0 (0.0%) | 26 (0.1%) |
| **Type of risk factor\*\*\*** |  |  |  |  |  |  |  |
| Any signs of danger or complication in current pregnancy - includes bleeding, convulsions, excessive vomiting, fainting, high fever, malpresentation of fetus and other (not defined) | 1502<br>(11.9%) | 755<br>(14.4%) | 2421<br>(23.1%) | 790<br>(9.4%) | 524 (7.4%) | 1 (1.1%) | 5993<br>(13.7%) |
| Maternal age at time of birth <16 or >40 | 288<br>(2.3%) | 186<br>(3.6%) | 442<br>(4.2%) | 238<br>(2.8%) | 342 (4.8%) | 14<br>(12.5%) | 1510<br>(3.4%) |
| Child is a multiple | 55 (0.4%) | 27<br>(0.5%) | 140<br>(1.3%) | 58 (0.7%) | 45 (0.6%) | 3 (2.7%) | 328<br>(0.7%) |
| Current baby very large or very small (proxy for fundal height not congruent with gestation) | 708<br>(5.6%) | 333<br>(6.4%) | 711<br>(6.8%) | 590<br>(7.0%) | 493 (7.0%) | 2 (1.6%) | 2838<br>(6.5%) |
| Ever had a stillbirth or neonatal death | 125<br>(1.0%) | 29<br>(0.6%) | 88<br>(0.8%) | 81 (1.0%) | 102 (1.4%) | 6 (5.5%) | 432<br>(1.0%) |
| Last birth a caesarean | 49 (3.7%) | 37<br>(5.5%) | 459<br>(47.8%) | 7 (0.5%) | 5 (0.4%) | 0 (0.0%) | 557<br>(10.2%) |
| Excessive vaginal bleeding at the time of birth of previous baby | 102<br>(0.8%) | 34<br>(0.7%) | 117<br>(1.1%) | 71 (0.8%) | 84 (1.2%) | 0 (0.0%) | 407<br>(0.9%) |

*\*\*In 2016 the World Health Organization released new antenatal care guidelines which increased its recommendations of a minimum of four to at least eight antenatal care visits with a health professional<sup>24</sup>.*
*\*\*\*Women can report more than one risk factor. Per cent is out of the total number of women with any risk factor.*

### Where did women with and without pregnancy risk give birth?

Figure 1 shows that across the 15 years of data, the proportion of women who gave birth in any facility (hospital, midwifery clinic, other clinic) has drastically increased from around 50% in 2002 to 84% by 2017. This was largely driven by the increase in hospital births which more than doubled from 16% in 2002 to 36% of all births in 2017 (see Figure 1). Conversely, the proportion of women who gave birth at home both with and without a health professional have more than halved over the same period. This shows a steady shift towards birthing in health facilities and away from giving birth at home over the 15-year period covered by the three waves of the survey.

**Figure 1:**
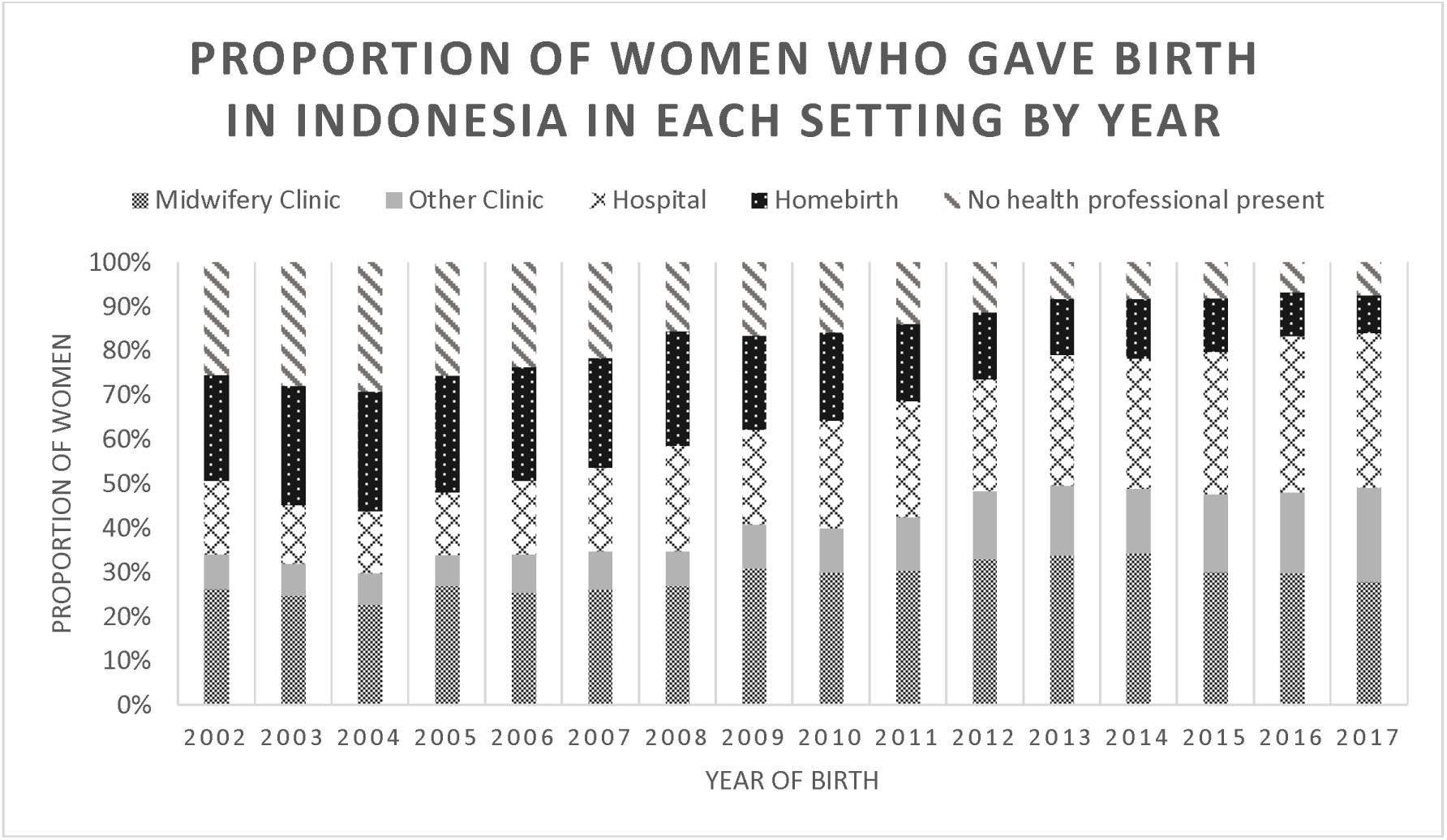
Proportions of women who gave birth to their most recent baby (born within five years of the survey) in different settings by year of birth 2002-2017 (weighted). here

Despite the overall shift away from giving birth at home over time, this does not appear to represent improvement in appropriate birth setting for those with risk factors. The proportion of women with risk factors who gave birth in each setting is shown in Figure 2. The figure demonstrates overall growth in reporting risk factors over time. It also shows that of the women who gave birth at home, the proportion who had risk factors has not gone down. Of the women who gave birth in hospital, a greater proportion reported pregnancy risk than in any other setting, and this went up from 0.32 in 2002 to 0.47 in 2017, averaging 0.35. This rise is not nearly as steep as the overall proportion of women who gave birth in hospital indicating that most of the doubling of hospital births has been from well women.

**Figure 2:**
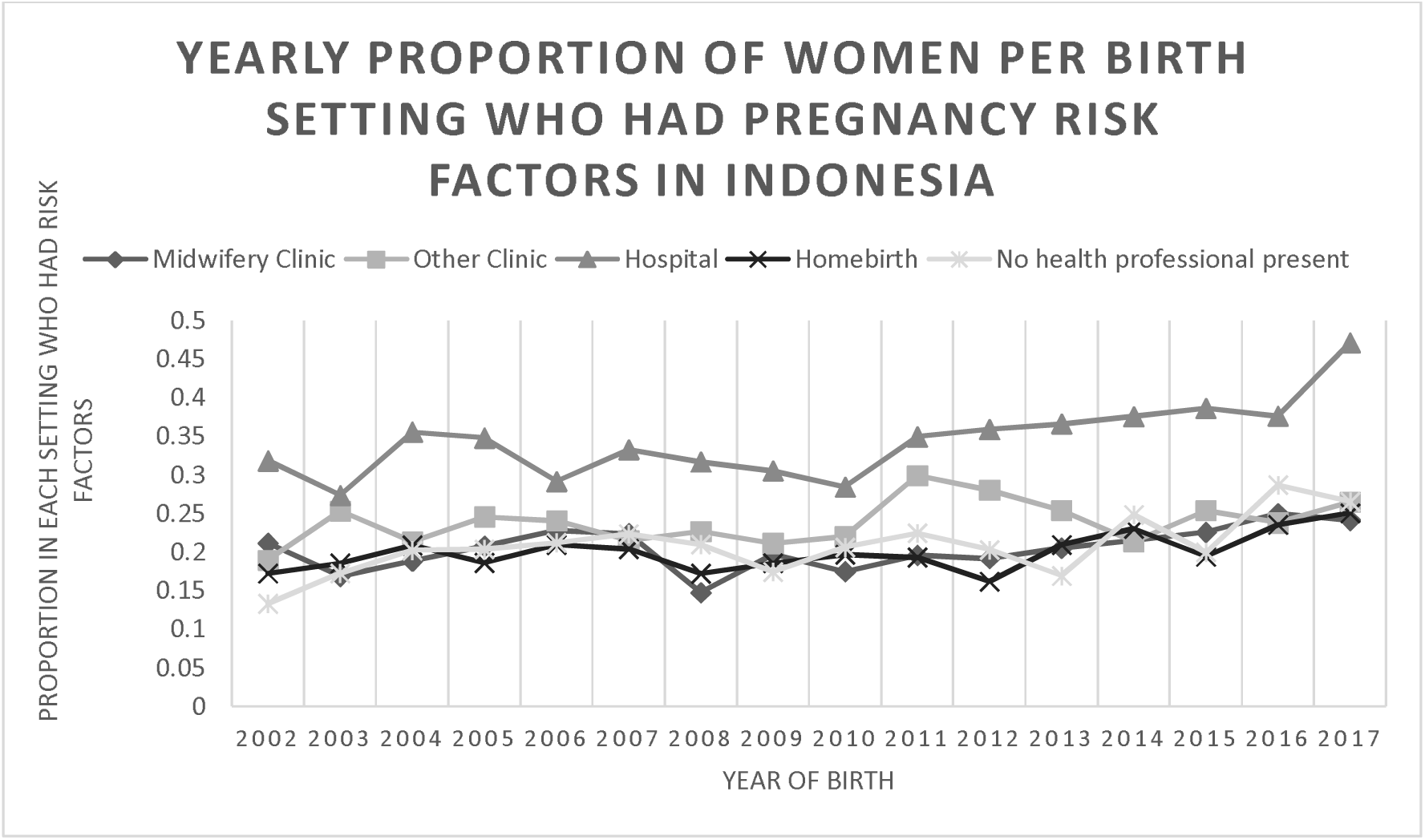
Yearly proportion of women per birth setting who had pregnancy risk factors in Indonesia over the years 2002- 2017

Three quarters of the women in the study population reported no risk factors. Less than 3% of the overall population reported more than one risk factor, with the remaining 21.6% of women reporting one risk factor. The most common setting for women with risk factors to give birth was in hospital. However, 1457, 1658 and 2587 women who had pregnancy risk factors gave birth respectively without a health professional present, at home, or in a midwifery clinic across the 15- year study period. Combined this equates to 13% of the total study population of women.

## Discussion

To our knowledge this is the first study of its kind to describe trends in place of birth by pregnancy-related risk factors in Indonesia, and this has rarely been done before outside of high-income countries. The large sample size and representative sampling are strengths of this study. Limitations include a risk of recall bias as women were asked retrospectively about their pregnancy risks and those who had an adverse outcome may have been more likely to report a risk in pregnancy.

The lack of a variable which asks women their intended place of birth is also a limitation of this study and of data from low and middle income countries generally. Intended place of birth would allow more nuanced and accurate assessment of the effectiveness of triaging and transfer of care as necessary.

Another limitation is that that not all risk factors are asked about in the Indonesian DHS dataset. In particular, many complications are only asked about in the most recent pregnancy, not previous pregnancies including history of convulsions, severe perineal tearing and assisted birth. Additionally, some of the questions ask for symptoms of a condition rather than actual diagnosis such as convulsions, swelling and headache as proxy for diagnosis of hypertensive disorders. Therefore, there is a lack of accuracy of risk factors when compared with a hospital-based dataset. This may result in an over or underestimation of risks in our study.

The first part of this paper identified variables in the Indonesian Demographic Health Survey Data which identified pregnancy risk factors from relevant referral guidelines. Using these datapoints, nearly one quarter of women reported retrospectively experiencing one or more risk factors in their pregnancy which would have resulted in recommendation of referral to a complex care facility. For comparison, smaller studies from other low and middle income countries have reported the incidence of high risk pregnancies to be 14.4%-25.8% ^24,25^. High income country studies report a transfer to hospital rate of 5-30% among planned homebirths ^26^. By these comparisons, our study reports a complexity proportion which could be described as reasonable.

Of the 24.4% of women who reported pregnancy-related risk factors, on average over half of them gave birth outside of hospital. This indicates a clear opportunity for task shifting in Indonesia that may optimise care for women and their babies.

Ninety-five per cent of women in Indonesia receive antenatal care from a health professional which is the ideal time to identify and recommend appropriate care for these risk factors. This study does not explain why, despite receiving antenatal care, many women gave birth in a setting not appropriate for their risk level. However, there is a wide body of literature on barriers to accessing of appropriate maternal healthcare most notably the delays model ^5^. Nearly 45% of women did not receive the WHO recommended 8 antenatal visits in their pregnancy which may have contributed to under recognition of risk factors. It is also possible that appropriate recommendations were not made despite care providers being aware of risk factors. Women may also have faced barriers to reaching care or chose for many reasons not to birth in recommended settings. Understanding these contributing factors and optimising referral pathways so that these women who are most likely to need complex obstetric or neonatal care have easy access at birth will likely result in reduced mortality and morbidity.

Additionally, our findings indicate that, though declining, on average over 60% of women who gave birth in a hospital did not report pregnancy complications. Encouraging these women without pregnancy risks to give birth in a more primary level of healthcare (such as a midwifery clinic or at home with a health professional) may mean more beds are available in referral level care for women who do need it. Midwives working in Primary Health Centres in Indonesia have reported difficulty in referrals for women being accepted by hospitals, which have been made worse by the demands of the Covid-19 pandemic ^27^. There is potential to improve the availability of referral beds if women without risk factors avoid hospitals, though more research is needed on this.

Additionally, supporting low risk women to birth outside of hospital settings may contribute to reducing the iatrogenic impact of overuse of interventions as well as reducing the costs on the health care system. While underuse of health care systems is a concern when a lack of necessary referral occurs, the opposite negative impact of the overuse of medicalisation at birth has been well described in the literature ^28^. A growing number of low and middle income countries are trending towards excessive, routine or inappropriate use of obstetric interventions such as episiotomies, routine continuous electronic fetal monitoring and non-medically indicated caesarean sections, labour induction and augmentations ^28,29^. Overusing these interventions increases both negative side-effects and sequelae for women and increases the costs and burden on the health system.

Shifting birth outside of hospitals for low risk women is frequently shown to reduce rates of a range of interventions ^26^.

### Conclusions

This paper highlights the crucial importance of provision of antenatal care and screening and recommending the appropriate place of birth for pregnant women. More research is needed into the outcomes of women who give birth in different settings both with and without risk factors to determine the safety of primary health care settings for birth outside of high income countries.

## Supporting information

Supplemental Table 1: STROBE checklist

## Data Availability

All data produced are available online with registration at

https://www.dhsprogram.com/data/available-datasets.cfm

## Acknowledgement

We, the authors Kai Hodgkin, Grace Joshy and Kamalini Lokuge agree that the article titled “Are women in Indonesia giving birth in the right setting for their level of pregnancy risk? Using Demographic Health Survey data to define pregnancy-related risk and assess appropriateness of place of birth” is our original work. The article has not received prior publication and is not under consideration for publication anywhere other than *Birth*. The authors have all seen and approved the manuscript being submitted.

This research is supported by an Australian Government Research Training Program (RTP) Scholarship provided to the first author. The Australian Government had no input into the topic, design or any other aspect of this research.

We declare no conflicts of interest.

## Ethics

This study received ethical approval from the chair of the Science and Medical Delegated Ethical Review Committee at the Australian National University on 2/10/2018. The protocol number is 2018/643.

## Notes

### Competing Interest Statement

The authors have declared no competing interest.

### Funding Statement

This study is supported by an Australian Government Research Training Program (RTP) Scholarship provided to the first author. The Australian Government had no input into the topic, design or any other aspect of this research.

## References

1. The World Bank. World Development Indicators. DataBank. Published 2020. Accessed November 30, 2020. https://databank.worldbank.org/reports.aspx?source=world-development-indicators

2. Koblinsky M, Moyer CA, Calvert C, et al. Quality maternity care for every woman, everywhere: a call to action. The Lancet. Published online September 2016. doi:10.1016/S0140-6736(16)31333-2

3. Renfrew MJ, McFadden A, Bastos MH, et al. Midwifery and quality care: findings from a new evidence-informed framework for maternal and newborn care. The Lancet. 2014;384(9948):1129–1145. doi:10.1016/S0140-6736(14)60789-3

4. Murray SF, Pearson SC. Maternity referral systems in developing countries: Current knowledge and future research needs. Social Science & Medicine. 2006;62(9):2205–2215. doi:10.1016/j.socscimed.2005.10.025

5. Thaddeus S, Maine D. Too far to walk: maternal mortality in context. Social science & medicine. 1994;38(8):1091–1110.

6. Gabrysch S, Campbell OM. Still too far to walk: Literature review of the determinants of delivery service use. BMC Pregnancy and Childbirth . 2009;9(1):34. doi:10.1186/1471-2393-9-34

7. Goodarzi B, Walker A, Holten L, et al. Towards a better understanding of risk selection in maternal and newborn care: A systematic scoping review. PLOS ONE. 2020;15(6):e0234252. doi:10.1371/journal.pone.0234252

8. Silver RM. Abnormal Placentation: Placenta Previa, Vasa Previa, and Placenta Accreta. Obstetrics & Gynecology. 2015;126(3):654–668. doi:10.1097/AOG.0000000000001005

9. National Institute for Health and Care Excellence. Intrapartum Care: Care of Healthy Women and Their Babies during Childbirth. version 2. National Institute for Health and Care Excellence; 2014. Accessed December 19, 2017. https://www.nice.org.uk/guidance/cg190/evidence/full-guideline-pdf-248734770

10. Bayrampour H, Lisonkova S, Tamana S, Wines J, Vedam S, Janssen P. Perinatal outcomes of planned home birth after cesarean and planned hospital vaginal birth after cesarean at term gestation in British Columbia, Canada: A retrospective population-based cohort study. Birth. 2021;48(3):301–308. doi:10.1111/birt.12539

11. Australian College of Midwives. National Midwifery Guidelines for Consultation and Referral . 4th ed. Australian College of Midwives; 2021. https://www.midwives.org.au/common/Uploaded%20files/_ADMIN-ACM/National-Midwifery-Guidelines-for-Consultation-and-Referral---4th-Edition_-(2021).pdf

12. Kementarian Kesehatan Republik Indonesia, World Health Organization, Perkupulan Obstetri dan Kinekologi Indonesia, Ikatan Bidan Indonesia. Buku Saku: Pelayanan Kesehatan Ibu Di Fasilitas Kesehatan Dasar Dan Rujukan . Kementerain Kesehatan Rebulik Indonesia; 2013. http://www.searo.who.int/indonesia/documents/976-602-235-265-5-buku-saku-pelayanan-kesehatan-ibu.pdf?ua=1

13. Hodgkin K, Joshy G, Browne J, Bartini I, Hull TH, Lokuge K. Outcomes by birth setting and caregiver for low risk women in Indonesia: a systematic literature review. Reproductive health. 2019;16(1):67.

14. Mgawadere F, Kana T, van den Broek N. Measuring maternal mortality: a systematic review of methods used to obtain estimates of the maternal mortality ratio (MMR) in low-and middle-income countries. British medical bulletin. 2017;121(1):121–134.

15. National Population and Family Planning Board (BKKBN), Statistics Indonesia (BPS), Ministry of Health (Kemenkes), ICF. Indonesia Demographic and Health Survey 2017 . BKKBN, BPS, Kemenkes, and ICF; 2018.

16. Mortality rate, neonatal (per 1,000 live births) - Indonesia, East Asia & Pacific. World Bank Open Data. Accessed September 10, 2023. https://data.worldbank.org

17. Deviany PE, Setel PW, Kalter HD, et al. Neonatal mortality in two districts in Indonesia: Findings from Neonatal Verbal and Social Autopsy (VASA). PLOS ONE. 2022;17(3):e0265032. doi:10.1371/journal.pone.0265032

18. von Elm E, Altman DG, Egger M, Pocock SJ, Gøtzsche PC, Vandenbroucke JP. The Strengthening the Reporting of Observational Studies in Epidemiology (STROBE) statement: guidelines for reporting observational studies. The Lancet. 2007;370(9596):1453–1457. doi:10.1016/S0140-6736(07)61602-X

19. Joint Committee on Reducing Maternal and Neonatal Mortality in Indonesia, Security Development, Policy and Global Affairs, National Research Council, Indonesian Academy of Sciences. The Indonesian Health Care System. National Academies Press (US); 2013. Accessed November 9, 2016. https://www.ncbi.nlm.nih.gov/books/NBK201708/

20. Bailey P, Lobis S, Fortney J, et al., eds. Monitoring Emergency Obstetric Care: A Handbook. World Health Organization; 2009.

21. StataCorp. Stata Statistical Software. Published online 2021.

22. World Health Organization, UNICEF. Pregnancy, childbirth, postpartum and newborn care: a guide for essential practice. Published online 2015.

23. World Health Organization. Global Programme on Evidence for Health Policy. Guidelines for WHO Guidelines. World Health Organization; 2003. Accessed November 10, 2022. https://apps.who.int/iris/handle/10665/68925

24. Rajbanshi S, Norhayati MN, Hazlina NHN. High-risk pregnancies and their association with severe maternal morbidity in Nepal: A prospective cohort study. PLOS ONE. 2020;15(12):e0244072. doi:10.1371/journal.pone.0244072

25. Ali A, Hora S, Agarwal G. A Cross Sectional Study on High Risk Pregnancy among Antenatal Women at Rural Primary Health Center in Eastern Part of Rajasthan. Published online 2019.

26. Zielinski R, Ackerson K, Kane Low L. Planned home birth: benefits, risks, and opportunities. Int J Womens Health . 2015;7:361–377. doi:10.2147/IJWH.S55561

27. Hazfiarini A, Zahroh RI, Akter S, Homer CSE, Bohren MA. Indonesian midwives’ perspectives on changes in the provision of maternity care during the COVID-19 pandemic: A qualitative study. Midwifery . 2022;108:103291. doi:10.1016/j.midw.2022.103291

28. Miller S, Abalos E, Chamillard M, et al. Beyond too little, too late and too much, too soon: a pathway towards evidence-based, respectful maternity care worldwide. The Lancet. Published online September 2016. doi:10.1016/S0140-6736(16)31472-6

29. Boerma T, Ronsmans C, Melesse DY, et al. Global epidemiology of use of and disparities in caesarean sections. The Lancet. 2018;392(10155):1341–1348. doi:10.1016/S0140-6736(18)31928-7

