## Supplemental Table 1: STROBE checklist for "Pregnancy-related risk and birth setting trends: Insights from Indonesian Demographic Health Survey data"

Appendix 1: STROBE Statement—Checklist of items that should be included in reports of ***cross-sectional studies***

|  | Item No. | Recommendation | Page No. | Relevant Text from Manuscript |
| --- | --- | --- | --- | --- |
| **Title and abstract** | 1 | (*a*) Indicate the study’s design with a commonly used term in the title or the abstract | Title page | Where are babies being born in Indonesia, and does it meet recommendations? Analysing cross-sectional Demographic Health Survey data to define pregnancy-related risk, time trends and appropriateness of birth setting |
|  |  | (*b*) Provide in the abstract an informative and balanced summary of what was done and what was found | 1 | Methods  Risk factors in pregnancy based on internationally relevant referral guidelines were identified from literature search and mapped to available indicators. Using self-reported data from three waves of the Indonesian Demographic Health Survey (2007, 2012, 2017), a representative survey of women aged 15-49 years, we examined receipt of maternity care, prevalence of pregnancy risk factors and time trends in birth setting, overall and by presence of risk factors.  Findings  In the weighted sample (n=43,846), one quarter of women reported pregnancy risk factors. From 2002-2017, numbers of births in hospitals have doubled and births at home have halved. However, the proportions of women with pregnancy risk in each setting remains largely unchanged. |
| Introduction | | |  |  |
| Background/rationale | 2 | Explain the scientific background and rationale for the investigation being reported | 3 | Despite the clear need to identify and triage care appropriately for women with risk factors in pregnancy, studies in low and middle income countries rarely, if at all, consider the presence of pregnancy or birth risks when assessing outcomes by birth settings 13. Therefore, there is no clear understanding of how well triaging to emergency obstetric health services on the basis of pregnancy risks is occurring in low and middle income countries. While this type of data is routinely collected in high income countries, many low and middle income countries do not have national pregnancy and birth datasets and therefore detail of pregnancy and birth care associated with complications is lacking 14. This kind of data is commonly only available at a hospital or health district level.  This article fills this gap by identifying in a population level dataset (Demographic Health Survey) pregnancy risk factors occurring prior to onset of labour that meet guidelines for referral to higher level services due to increased risk of adverse outcomes. We then describe where these women with and without pregnancy risks gave birth which indicates whether appropriate triaging has occurred. |
| Objectives | 3 | State specific objectives, including any prespecified hypotheses | 4 | Aim  We developed two specific aims for this project:  1. Outline a set of indicators for use in Indonesia which define women with pregnancies at risk of obstetric complications during birth, and well women with lower risk of obstetric complications during birth utilising Indonesian Demographic Health survey data variables.  2. Determine trends in birth settings overall, and for women with and without pregnancy risk in Indonesia. |
| Methods | | |  |  |
| Study design | 4 | Present key elements of study design early in the paper | 4, 5, 6 | Retrospective, cross-sectional Demographic Health Survey (DHS) data from Indonesia was utilised for this investigation including the most recent three waves of the survey: 2007, 2012, 2017.  …we searched the internet for relevant care guidelines which included a list of risk factors which would result in the recommendation of referral to a setting with emergency obstetric (and/or neonatal) care and facilities. These risk factors were then compiled and organised by domain (maternal, neonatal and obstetric history). The Indonesian DHS dataset was then searched for an equivalent or proxy variable for each of these factors which could be utilised in this analysis.  Weighted proportions were estimated, applying DHS-provided weights for each wave. In this population the weighting includes correcting for non-response and oversampling of women in rural households. Initial cross-tabulations were undertaken ensuring data accuracy. |
| Setting | 5 | Describe the setting, locations, and relevant dates, including periods of recruitment, exposure, follow-up, and data collection | 4-5 | Data  Retrospective, cross-sectional Demographic Health Survey (DHS) data from Indonesia was utilised for this investigation including the most recent three waves of the survey: 2007, 2012, 2017. DHS is a USAid funded international initiative which focuses on collecting high quality, representative survey data in conjunction with lower and middle income countries. Each wave of the survey involves a centrally developed questionnaire which is then modified, translated and administered by each local country involved. The dataset is de-identified, coded, cleaned and released publicly on the DHS website for those who register. The process of participant selection, data collection and documentation is well documented elsewhere 15.  Population  The population was defined as women aged 15-49, interviewed in the 2007, 2012 or 2017 Indonesian DHS waves, who had given birth to their most recent baby within five years of the survey. In 2007, the survey was only conducted among married women, while in 2012 and 2017, unmarried women were also included. |
| Participants | 6 | (*a*) Give the eligibility criteria, and the sources and methods of selection of participants | 4 | As this is a secondary analysis, reference is made to sources which describe in detail the method of selection of participants and their eligibility criteria:  National Population and Family Planning Board (BKKBN), Statistics Indonesia (BPS), Ministry of Health (Kemenkes), ICF. Indonesia Demographic and Health Survey 2017. Jakarta, Indonesia: BKKBN, BPS, Kemenkes, and ICF, 2018. |
| Variables | 7 | Clearly define all outcomes, exposures, predictors, potential confounders, and effect modifiers. Give diagnostic criteria, if applicable | 5 | Defining increased risk of requiring obstetric intervention  We aimed to identify a definition of increased risk of obstetric complications in pregnancy which is applicable in Indonesia and from an international organisation such as the World Health Organisation. To do this, we searched the internet for relevant care guidelines which included a list of risk factors which would result in the recommendation of referral to a setting with emergency obstetric (and/or neonatal) care and facilities. These risk factors were then compiled and organised by domain (maternal, neonatal and obstetric history). The Indonesian DHS dataset was then searched for an equivalent or proxy variable for each of these factors which could be utilised in this analysis.  We then undertook an analysis of place of birth by the number of risk factors a woman self-reported. This was based on an assumption that the greater the number of risk factors a woman experiences in her pregnancy the greater the risk of complications at birth, and therefore the more important it would be that the woman gives birth in a setting with complex care available (tier two from the Renfrew Framework 3).  Categorising birth settings  Indonesia has a range of types of health facilities where women birth 19. Across each of the three waves of the Indonesian DHS these were defined differently so existing categories could not be used. Additionally, some categories contained too few births to result in useful findings. For the purposes of this study, birth settings were grouped into categories aiming to approximate the level of tier two care 20 available: Hospitals (including public, private and maternity hospitals); other clinics (these include Puskesmas, Posyandu, PolIndes, private nurse, GP and obstetric practices); midwifery clinics (including government funded and private practice clinics); and homebirth (including another person’s home) where a health professional (mostly a midwife) is present. The final category is all births not attended by a health professional (many were attended by a dukun bayi or traditional birth attendant), 98 per cent of which occur at home and also includes most births in the ‘other’ category which (while unclear) likely involves babies born en route to a health facility or at un- or under-staffed clinics.  While the categories do not explicitly equate to the types of services available in each setting, we assume that at the majority of both public and private hospitals, Comprehensive Emergency Obstetric and Neonatal Care (CEmONC) facilities will be available while clinics (including midwifery clinics) might have access to some, if not all Basic Emergency Obstetric and Neonatal Care requirements (BEmONC). BEmONC facilities are usually able to provide life-saving medications, intravenous therapy, basic neonatal resuscitation and assisted birth 20. CEmONC facilities can provide the same as BEmONC in addition to emergency surgery (caesarean section) and specialist obstetric and neonatal care. Homes are unlikely to have any of these, though if health professionals are in attendance, they likely bring assessment tools, sterile birth kits, some medications, intravenous therapy and basic resuscitation equipment. |
| Data sources/ measurement | 8* | For each variable of interest, give sources of data and details of methods of assessment (measurement). Describe comparability of assessment methods if there is more than one group | 5 | All variables used in this analysis are based on self-report of the mothers surveyed. |
| Bias | 9 | Describe any efforts to address potential sources of bias | 6 | Weighted proportions were estimated, applying DHS-provided weights for each wave. In this population the weighting includes correcting for non-response and oversampling of women in rural households.  Recall bias is acknowledged as a potential limitation in the discussion. |
| Study size | 10 | Explain how the study size was arrived at | 5 | The population was defined as women aged 15-49, interviewed in the 2007, 2012 or 2017 Indonesian DHS waves, who had given birth to their most recent baby within five years of the survey. In 2007, the survey was only conducted among married women, while in 2012 and 2017, unmarried women were also included. |
| Quantitative variables | 11 | Explain how quantitative variables were handled in the analyses. If applicable, describe which groupings were chosen and why |  | N/A |
| Statistical methods | 12 | (*a*) Describe all statistical methods, including those used to control for confounding | 6 | Weighted proportions were estimated, applying DHS-provided weights for each wave. |
|  |  | (*b*) Describe any methods used to examine subgroups and interactions |  | N/A |
|  |  | (*c*) Explain how missing data were addressed | 7 | Table 2 describes the demographics of women included, including missing items where applicable. |
|  |  | (*d*) If applicable, describe analytical methods taking account of sampling strategy |  | N/A |
|  |  | (*e*) Describe any sensitivity analyses |  | N/A |
| Results | | |  |  |
| Participants | 13* | (a) Report numbers of individuals at each stage of study—eg numbers potentially eligible, examined for eligibility, confirmed eligible, included in the study, completing follow-up, and analysed |  | N/A – as secondary analysis recruitment is not part of this study. |
|  |  | (b) Give reasons for non-participation at each stage |  | N/A |
|  |  | (c) Consider use of a flow diagram |  | N/A |
| Descriptive data | 14* | (a) Give characteristics of study participants (eg demographic, clinical, social) and information on exposures and potential confounders |  | See table 2 |
|  |  | (b) Indicate number of participants with missing data for each variable of interest |  | Table 2 |
| Outcome data | 15* | Report numbers of outcome events or summary measures |  | Table 3 |
| Main results | 16 | (*a*) Give unadjusted estimates and, if applicable, confounder-adjusted estimates and their precision (eg, 95% confidence interval). Make clear which confounders were adjusted for and why they were included |  | N/A |
|  |  | (*b*) Report category boundaries when continuous variables were categorized |  | N/A |
|  |  | (*c*) If relevant, consider translating estimates of relative risk into absolute risk for a meaningful time period |  | N/A |
| Other analyses | 17 | Report other analyses done—eg analyses of subgroups and interactions, and sensitivity analyses |  | N/A |
| Discussion | | |  |  |
| Key results | 18 | Summarise key results with reference to study objectives | 9 | The first part of this paper identified variables in the Indonesian Demographic Health Survey Data which identified pregnancy risk factors from relevant referral guidelines. Using these datapoints, nearly one quarter of women reported retrospectively experiencing one or more risk factors in their pregnancy which would have resulted in recommendation of referral to a complex care facility.  The second aim was to determine trends in birth settings, with reference to those who have risk factors. Of the 24.4% of women who reported pregnancy-related risk factors, on average over half of them gave birth outside of hospital. This indicates a clear opportunity for task shifting in Indonesia that may optimise care for women and their babies. |
| Limitations | 19 | Discuss limitations of the study, taking into account sources of potential bias or imprecision. Discuss both direction and magnitude of any potential bias | 8-9 | Limitations include a risk of recall bias as women were asked retrospectively about their pregnancy risks and those who had an adverse outcome may have been more likely to report a risk in pregnancy.  The lack of a variable which asks women their intended place of birth is also a limitation of this study and of data from low and middle income countries generally. Intended place of birth would allow more nuanced and accurate assessment of the effectiveness of triaging and transfer of care as necessary.  Another limitation is that that not all risk factors are asked about in the Indonesian DHS dataset. In particular, many risk factors are only asked about in the most recent pregnancy, not previous pregnancies including history of convulsions, severe perineal tearing and assisted birth. Additionally, some of the questions ask for symptoms of a condition rather than actual diagnosis such as convulsions, swelling and headache as proxy for diagnosis of hypertensive disorders. Therefore, there is a lack of accuracy of risk factors when compared with a hospital-based dataset. This may result in an over or underestimation of risks in our study. |
| Interpretation | 20 | Give a cautious overall interpretation of results considering objectives, limitations, multiplicity of analyses, results from similar studies, and other relevant evidence | 10 | This paper highlights the crucial importance of provision of antenatal care and screening and recommending the appropriate place of birth for pregnant women. It is clear that not all women are giving birth in a setting that can best provide appropriate care for their level of risk in pregnancy. More research is needed into the outcomes of women who give birth in different settings both with and without risk factors to determine the safety of primary health care settings for birth outside of high income countries. More work is needed also to identify and overcome the barriers to women in need accessing appropriate care at birth. |
| Generalisability | 21 | Discuss the generalisability (external validity) of the study results | 10 | This paper targets Indonesian data and therefore the findings may not be applicable to other countries. However, the implications that birth setting should be analysed in the contexts of pregnancy complications is an important one to other low resource settings. |
| Other information | | |  |  |
| Funding | 22 | Give the source of funding and the role of the funders for the present study and, if applicable, for the original study on which the present article is based | 11 | This research is/was supported by an Australian Government Research Training Program (RTP) Scholarship held by first author. The scholarship funder had no contribution to the topic, data analysis or interpretation. The funder has no right to approve or otherwise this publication. The authors declare no conflicts of interest. |

*Give information separately for exposed and unexposed groups.

**Note:** An Explanation and Elaboration article discusses each checklist item and gives methodological background and published examples of transparent reporting. The STROBE checklist is best used in conjunction with this article (freely available on the Web sites of PLoS Medicine at http://www.plosmedicine.org/, Annals of Internal Medicine at http://www.annals.org/, and Epidemiology at http://www.epidem.com/). Information on the STROBE Initiative is available at www.strobe-statement.org.
